# *FGF14* repeat length and mosaic interruptions: modifiers of SCA27B?

**DOI:** 10.1101/2024.11.21.24317532

**Authors:** Joshua Laß, Mirja Thomsen, Max Borsche, Theresa Lüth, Julia C. Prietzsche, Susen Schaake, Andona Milovanović, Hannah Macpherson, Emil K. Gustavsson, Paula Saffie Awad, Nataša Dragašević-Mišković, Björn-Hergen Laabs, Inke R. König, Ana Westenberger, Christopher E. Pearson, Norbert Brüggemann, Christine Klein, Joanne Trinh

## Abstract

Deep intronic *FGF14* repeat expansions have been identified as a frequent genetic cause of late-onset cerebellar ataxias, explaining up to 30% of patients. Interruptions between repeats have previously been identified to impact the penetrance in other repeat expansion disorders. Repeat interruptions within *FGF14* have yet to be characterized in detail.

We utilized long-range PCR, Sanger sequencing, repeat-primed PCR, Nanopore, and PacBio sequencing to distinguish the repeat motifs, mosaicism, and number of repeat interruptions present in *FGF14*-related ataxia patients and unaffected individuals.

A total of 304 patients with late-onset ataxia and 190 unaffected individuals were previously screened for repeat expansions in the *FGF14* gene by long-range PCR, identifying 37 individuals with expanded repeat lengths (≥250 repeats). These, along with three newly identified expansion carriers were included in the present study, and advanced genetic methods were applied to investigate the repeat composition in 27 patients and 13 unaffected individuals. The expansions, based on Nanopore data, ranged from 236 to 486 repeats (SD = 60), with 20 individuals showing repeat interruptions, including complex motifs such as *<u>GAG</u>*, GAA*<u>GGA</u>*, GAAGAA*<u>A</u>*GAA, GAAA*<u>A</u>*GAAGAA*<u>G</u>*GAAGAA*<u>G</u>*GAA, GAAA*<u>A</u>*GAAGAA*<u>G</u>*GAA, and *<u>GCA</u>*GAAGAAGAAGAA. We calculated the longest pure GAA length from the long-read data for all 40 individuals. When comparing the pure GAA tract between patients and unaffected individuals, clusters were apparent based on greater or less than 200 repeats. Five ataxia patients with interruptions still had a remaining pure GAA expansion >200. We observed an association of the pure GAA length with age at onset (*p*=0.016, *R*^2^=0.256). Somatically-incurred mosaic divergent repeat interruptions were discovered that affect motif length and sequence (mDRILS), which varied in number and mosaicism (frequency: 0.37-0.93). The mDRILS correlated with pure GAA length (*p*=0.022, *R*^2^=0.334), with a higher mosaic frequency of interruptions in unaffected individuals compared to patients (unaffected: 0.90; patients: 0.67; *p*=0.009).

We demonstrate that i) long-read sequencing is required to detect complex repeat interruptions accurately; ii) repeat interruptions in *FGF14* are mosaic, have various lengths and start positions in the repeat tract, and can thereby be annotated as mDRILS, which iii) enabled us to establish a categorization based on remaining pure GAA repeats quantifying the impact of mDRILS on pathogenicity or age at onset, dependent on the interruption length and position, with high accuracy. Finally, we iv) provide evidence that mosaicism stabilizes pure GAA repeats in interrupted *FGF14* repeat expansions.

## Introduction

Genetic cerebellar ataxias are commonly associated with tandem repeat expansions. The recently identified deep intronic repeat expansions of (GAA)_n_ in the *FGF14* (Fibroblast Growth Factor) gene cause SCA27B.^1,2^ The expansion is located in the first intron and leads to a reduction of *FGF14* expression, similar to the GAA expansion of Friedrich’s ataxia.^3^ In SCA27B, a threshold of ≥250 repeats is considered disease-causing, whereby expansions between 250 and 300 repeats are likely pathogenic with reduced penetrance, and expansions with ≥300 repeats are fully penetrant.^1,2^ Different studies suggested repeat interruptions in *FGF14* to be non-pathogenic.^2,4–7^ However, evidence for interruption non-pathogenicity relates mainly to family studies thus far, and more in-depth analysis of the specific repeat expansion sequence, motif and interruption length has yet to be performed. One large study used Nanopore sequencing in a subset of individuals and found a similar frequency of GAA*<u>GGA</u>* interruptions in patients and controls, where they concluded that the interruption was non-pathogenic.^6^ However, interruptions have not been further characterized in terms of interruption length, position, and mosaicism. Finally, the role of shorter repeat interruptions and the impact of their position within the repeat has yet to be deciphered in detail.

Long-read sequencing has revealed an increased appreciation of the number of loci with expanded repeats, the sequence motifs, and their purity.^8,9^ While many repeat expansion disorders are now characterized, with known *cis*-elements flanking the unstable repeat, including *FGF14* repeat tract purity, modifiers of disease expression are largely unknown.^10,11^ The hexanucleotide repeat relevant for X-linked dystonia-parkinsonism (XDP) consists of the hexanucleotide (*AGAGGG*)_n_ sequence repeated 30 to 55 times and is a strong genetic modifier of age at onset (AAO).^12,13^ A novel mosaic repeat motif pattern that deviates from the known hexanucleotide repeat motif, both in motif length and sequence (mDRILS), modifies repeat stability in XDP.^14^ This genetic association in XDP demonstrates the importance of somatic mosaic genotypes and the biological plausibility of multiple germline and somatic modifiers, which may collectively contribute to repeat instability. These variations may remain undetected without assessment of single molecules. Data on the correlation between repeat length and age at onset varies between *FGF14* studies. Even studies with large sample sizes could not consistently demonstrate such an association, which contradicts with the general understanding of repeat expansion disorders.^15–17^ Thus, in-depth genetic methods might shed new light on this relation, which is of importance for clinical care and patient counseling.

Our study aimed to delineate the repeat tract sequence, mosaicism, and number of repeat interruptions in *FGF14*-related late-onset cerebellar ataxia patients and unaffected individuals (Fig. 1). Our findings show that: i) long-read sequencing is required to detect complex repeat interruptions accurately; ii) repeat interruptions in *FGF14* are mosaic, have various lengths and start positions in the repeat tract, and can thereby be annotated as mDRILS, which iii) enabled us to establish a categorization based on remaining pure GAA repeats quantifying the impact of mDRILS on pathogenicity or age at onset, dependent on the interruption length and position, with high accuracy. Finally, we iv) provide evidence that mosaicism stabilizes pure GAA repeats in interrupted *FGF14* repeat expansions.

**Figure 1.**
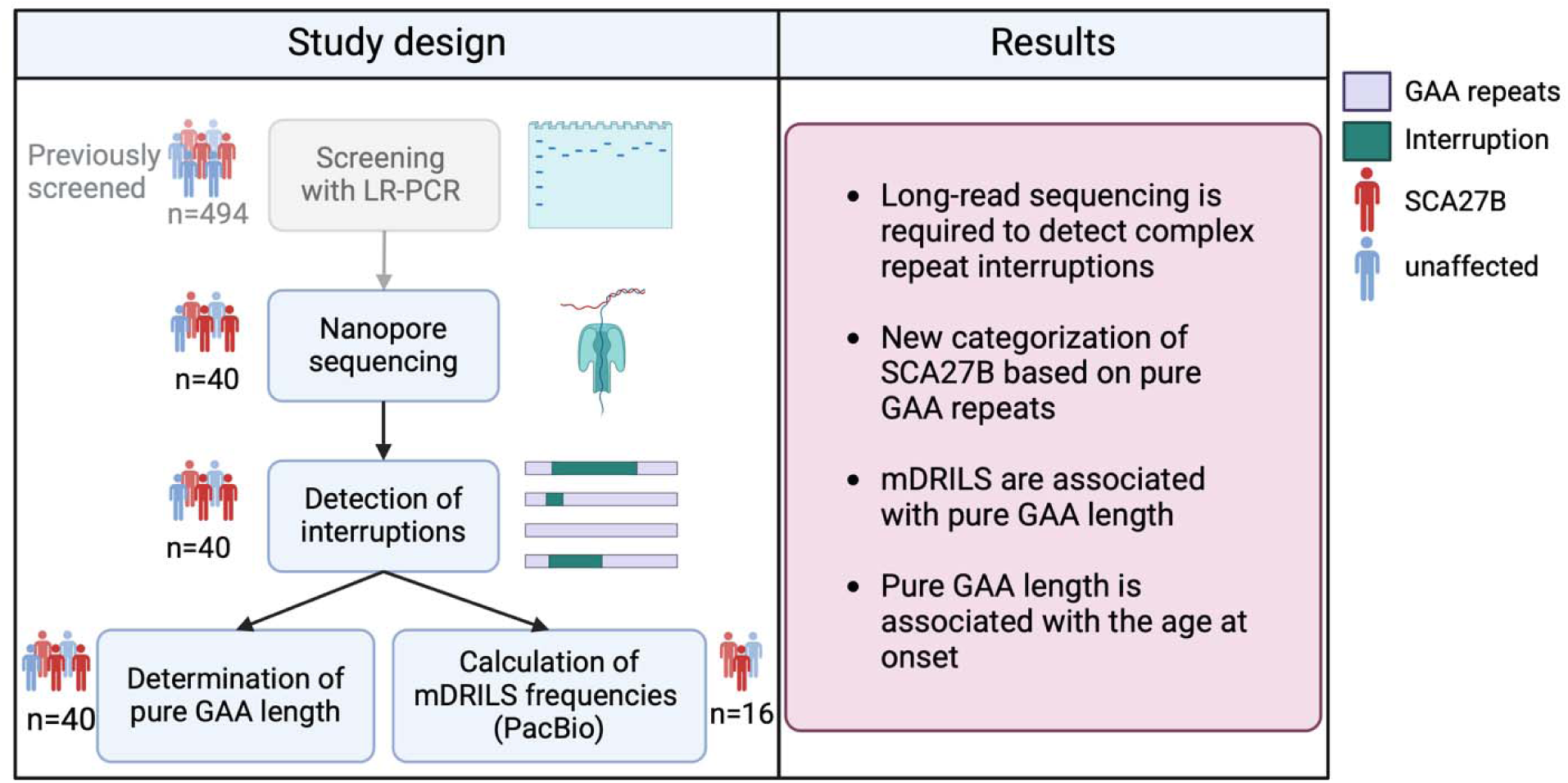
Overview of the study design. Initial screening of *FGF14* repeat expansions (≥250 repeats) was previously performed with long-range PCR.^1,17–19^ In the present study, individuals with detected expansions were further analyzed using long-read sequencing to detect interruptions, pure GAA tract length, and somatic mosaicism. Legend: LR-PCR = long-range PCR, PacBio = PacBio sequencing, mDRILS = mosaic divergent repeat interruption affecting length and sequence. Created in BioRender. Laß, J. (2025) https://BioRender.com/a67g089.

## Methods

### Participant recruitment

The individuals analyzed in greater detail in this study (n=40) were selected from previous (n=37) and ongoing (n=3) screening efforts for *FGF14* repeat expansions conducted at the Institute of Neurogenetics, Lübeck, Germany (Fig. 1). These screening efforts included i) patients with ataxia of unknown genetic cause (134 patients from Germany recruited at the tertiary referral centers for ataxia and vertigo at the Department of Neurology, University of Lübeck, Germany,^1,18^ 167 patients from Serbia recruited at the Department for Neurodegenerative Diseases and Movement Disorders, Neurology Clinic at the University Clinical Center of Serbia, Belgrade, Serbia,^19^ and two patients and one unaffected member of a Chilean family recruited at a movement disorders center in Santiago, Chile.^20^); and ii) 190 unaffected individuals from Germany.^1^ Control participants were recruited within the framework of large cross-sectional or longitudinal studies independent from ataxia research dealing with Parkinson’s disease or eating behavior at the Institute of Neurogenetics, Lübeck, Germany.^21,22^ All control participants underwent structured neurological examination and were free of signs of neurological disease at the time of blood taking, which was set as age at examination.

The inclusion criterion for ataxia patients was the presence of progressive cerebellar ataxia of unknown cause. Patients with secondary forms of ataxia (lesional, toxic, inflammatory, or paraneoplastic) and known repeat-expansion SCAs (SCA1, 2, 3, 6, and 17) were excluded. The study was approved by the local ethics committees at the University of Lübeck, Germany, the Medical Faculty of the University of Belgrade, Serbia, and CETRAM (Centro de Estudios de Transtornos del Movimiento), Chile. All patients gave written informed consent prior to inclusion in the study, which was performed according to the Declaration of Helsinki. After the collection of peripheral blood samples, genomic DNA was extracted using the QIAamp DNA Blood Mini Kit (Qiagen) according to the manufacturer’s instructions.

### Long-range PCR

Long-range PCR was performed to amplify the GAA repeat region of *FGF14* using flanking primers FGF14-F (5’-CAGTTCCTGCCCACATAGAGC-3’) and FGF14-R (5’-AGCAATCGTCAGTCAGTGTAAGC-3’), as previously described.^1^ The predicted product is 315 bp long and includes 50 GAA repeats based on the hg38 reference. A 25 µL PCR reaction was set up using the Platinum SuperFi II Master Mix (Thermo Fisher Scientific), 5% DMSO, 0.5 µM forward and reverse primers, and 100 ng of genomic DNA, with amplification performed using a touchdown PCR protocol (Supplementary Table 1). PCR products were visualized using agarose gel electrophoresis. For fragment analysis, an M13F-tail (CACGACGTTGTAAAACGAC) was attached to the forward primer, and a third FAM-labeled primer (FAM-M13F) was added to analyze products by capillary electrophoresis on a Genetic Analyzer 3500XL (Applied Biosystems). Fragment sizes were determined using GeneMapper software (Applied Biosystems) with a GeneScan™ 1200 LIZ™ Size Standard. For samples with repeat lengths greater than 250, corresponding to products with a size greater than ∼900 bp, repeat-primed PCR (RP-PCR), Sanger sequencing, Nanopore, and PacBio sequencing were conducted.

### Repeat-primed PCR (RP-PCR)

RP-PCR was used to analyze the repeat composition and test for possible interruptions of the (GAA)n repeat, following a protocol and primers adapted from a previous publication.^1^ The design included a locus-specific primer (FGF14-R), a repeat-containing primer with an M13F-tail designed to amplify the GAA/TCC or GAAGGA/CTTCCT motif (FGF14-RP-GAA: 5’-M13F-CTTCTTCTTCTTCTTCTTCTT-3’; FGF14-RP-GAAGGA: 5’-M13F-TCCTTCTCCTTCTCCTTC-3’), and a FAM-labeled M13-primer (FAM-M13F). Cycling conditions are displayed in Supplementary Table 2. The products were analyzed on a Genetic Analyzer 3500XL, producing a ladder-like pattern indicative of repeat expansions of the respective motif.

### Nanopore sequencing

A Nanopore sequencing workflow was used to analyze the repeat expansion in *FGF14*. The approach involved PCR amplification of the repeat tract, utilizing the long-range PCR products (see above) as an input.

To sequence all individuals on a single R10.4.1 flow cell, the long-range PCR products were multiplexed using the Native Barcoding Kit NBD114-96 (ONT). A 200 fmol input of the amplified PCR product was used. Library preparation was performed with the SQK-LSK114 kit

(ONT). The final library was subsequently loaded onto an R10.4.1 flow cell (FLO-MIN114) and sequenced on a GridION platform as previously described.^23^

### Sanger sequencing

Sanger sequencing was performed on the long-range PCR products to confirm their specificity and the repeat motif sequence. Sequencing reactions were prepared using the BigDye Terminator v3.1 Cycle Sequencing Kit (Applied Biosystems) with primers FGF14-F and FGF14-R. The sequencing conditions included 25 cycles of 96 °C for ten seconds, 55 °C for five seconds, and 60 °C for three minutes. Sequencing products were purified using sodium acetate precipitation and analyzed on a Genetic Analyzer 3500XL (Applied Biosystems).

### Pacbio sequencing

For each sample, a 1.3X cleanup was performed using PacBio SMRTbell beads. Quality control checks were conducted using the Qubit 1x dsDNA HS kit (Thermo Fisher) for concentration and the Femto Pulse with the Genomic DNA 165kb Kit (Agilent) for fragment analysis. Samples were then processed using an adapted version of the standard PacBio multiplexed amplicon library protocol (102-359-000) and the SMRTbell Prep Kit 3.0. A final concentration of 78.38 fmol of each sample was added, and reaction volumes were divided by six for End Repair and A-tailing and by 5.4 for Adapter Ligation. During ligation, samples were barcoded with unique SMRTbell adapters, and an additional step of incubation at 65 °C for ten minutes was added before the four °C hold. Samples were pooled before a 1.2X Pronex bead clean and elution in 40 μL. Nuclease treatment was performed in a total volume of 50 μL at 37 °C for 15 min and then held at four °C. A second 1.2X Pronex bead cleanup was carried out with final elution in 15 μL. The final library underwent quality assessment using the Qubit and Femto Pulse. Sequencing was carried out as a standard amplicon library on the Sequel IIe using binding kit 3.2. The library was loaded at 70 pM with a movie time of 30 hours.

### Bioinformatic analysis

For Nanopore sequencing, base-calling was performed using Dorado’s (version 7.2.13) super accuracy model (dna_r10.4.1_e8.2_400bps_sup@v4.3.0). Read quality was analyzed with the Nanostat software (version 1.5.0). For PacBio sequencing, the BAM files were demultiplexed with the software Lima (version 2.9.0) and converted to FASTQ files using Samtools (version 1.15). For both Nanopore and PacBio sequencing, Minimap2 (version 2.22) was used to align the reads to the reference sequence with parameters for long Nanopore sequencing reads.^24^ SAM-to-BAM conversion and BAM file handling were conducted with the Samtools software (version 1.15).^25^ The next step was the sorting and indexing of the reads with Samtools.

The trinucleotide repeats were analyzed with the “Noise-Cancelling Repeat Finder” (NCRF, version 1.01.02).^26^ Only reads with a maximum noise of 80% and a minimum of 15 detected repeat units were included in the analysis. A minimum threshold of 200 repeats was set to filter for the long allele. A maximum threshold of 100 repeats was applied to assign reads to the short allele. The repeat length was determined with the median repeat length of all reads. The detection of interruptions and their frequency were done with the summary output in R, as previously described.^23^ The interruption frequency was then calculated by dividing the number of reads with interruption by the total number of reads for that individual. The scripts and reference file are provided at: https://github.com/joshua21997/FGF14-repeat-expansion.

### Statistical analysis

The graphical representation and statistical analyses were performed in R (version 4.3.0) and Biorender. Visualization was done with the ggplot2 package (version 3.4.4). Mann-Whitney U-tests were performed for pairwise comparisons between patients and unaffected individuals, with the significance level set to α = 0.05 for the Mann-Whitney U-tests. The adjusted significance level for multiple testing based on Bonferroni is α = 0.013. To compare the different methods, Bland-Altman plots were used. Additionally, correlation analysis using a linear regression model implemented with the lm-function was performed.

## Data availability

The data that support the findings of this study are available from the corresponding author, upon reasonable request.

## Results

### Long-read sequencing can robustly detect *FGF14* repeat length

Nanopore sequencing on the above-described 40 individuals with *FGF14* repeat expansions revealed a read length of the long allele at a median of 1018 bp (IQR:899-1112 bp), and the median q-score was 14.6 (IQR:14.2-15.4). The detected repeat number with Nanopore sequencing ranged from 236 to 486, and the median repeat length was 309 (IQR:276-373) (Supplementary Table 3). The fragment analysis detected comparable repeat lengths for the long allele (median repeat length: 324, IQR:288-376) (Fig. 2A). The Bland-Altman analysis between Nanopore sequencing and fragment analysis showed a small bias, and three of 40 individuals were out of the limits of agreement (Fig. 2B).

**Figure 2.**
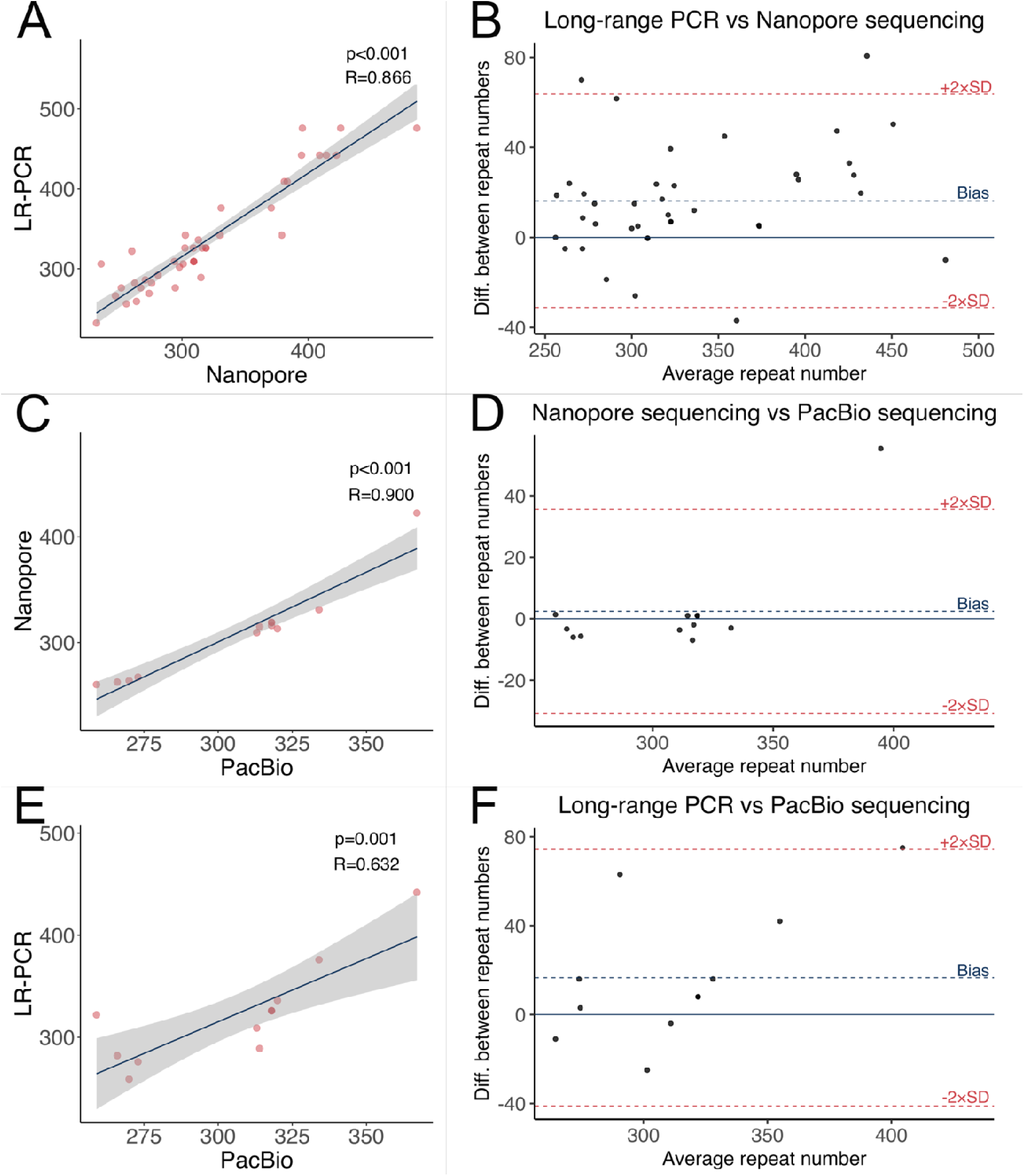
Comparison of the repeat length between different methods. **(A)** Correlation between LR-PCR and Nanopore. **(B)** Bland Altman Plots between LR-PCR and Nanopore **(C)** Correlation between Nanopore and PacBio. **(D)** Bland Altman Plots between Nanopore and PacBio. **(E)** Correlation between LR-PCR and PacBio. **(F)** Bland Altman Plots between LR-PCR and PacBio. A linear regression model was used for statistical analysis. Legend: LR-PCR = long-range PCR, Nanopore = Oxford Nanopore Technology sequencing, PacBio = PacBio sequencing.

PacBio sequencing was performed on 16/40 individuals with the *FGF14* repeat expansion as a second validation of the repeat length. The median q-score of the PacBio sequencing was 38.7 (IQR:37.7-41.5), and the median read length was 1162 bp (IQR:1108-2310 bp). The median repeat number of the long allele with PacBio sequencing was 318 (IQR:273-342).

Comparing the repeat tract length, the PacBio sequencing results were concordant with the Nanopore sequencing results and with the fragment analysis results (one of 16 individuals was outside the limits of agreement) (Fig. 2C-F).

### The pure GAA tract predicts disease manifestation and age at onset

Next, we assessed the pure GAA tract length, without interruptions, in patients and unaffected individuals. The range of the pure GAA tract was 11 to 486 repeat units across these individuals. For individuals with interruptions, the longest GAA tract was used for further analyses. The comparison between patients and unaffected individuals resulted in the identification of four distinct clusters (Fig. 3A and 4A). Recent literature has identified SCA27B patients with a lower repeat number than 250.^6,27^ We observed a gap between 66 to 226 pure GAA repeats (Fig. 4A). Therefore, we used a threshold of 200 pure GAA repeat units, albeit in a suggestive manner. The first group consisted of affected patients with a pure GAA length of >200 repeat units and diagnosed with SCA27B (affected, Fig. 3A and 4A indicated in red). In the second group were unaffected individuals with a pure GAA tract >200, indicating pre-manifesting carrier status (pre-manifesting, Fig. 3A and 4A indicated in yellow). Regarding AAO, the median AAO of SCA27B patients is reported to be 60 years (range 21-87 years)^28^, which we used as a supposed threshold for the categorization of affected and pre-manifesting individuals in Figure 3A. Of note, these six individuals had an age at examination of 6, 35, 50, 51, 53, and 80 years, respectively. The third group consisted of unaffected individuals with a pure GAA length <200 and no diagnosis (unaffected, Fig. 3A and 4A indicated in blue). The last group consisted of patients with a pure GAA tract <200 repeat units (other ataxia, Fig. 3A and 4A indicated in green). Next, we tested the relationship between the pure GAA lengths and AAO (Figure 3B). There was an association between GAA repeat length and AAO (*p*=0.016, *R*^2^=0.256) (Fig. 4B) in patients with a pure GAA tract >250 repeat units.

**Figure 3.**
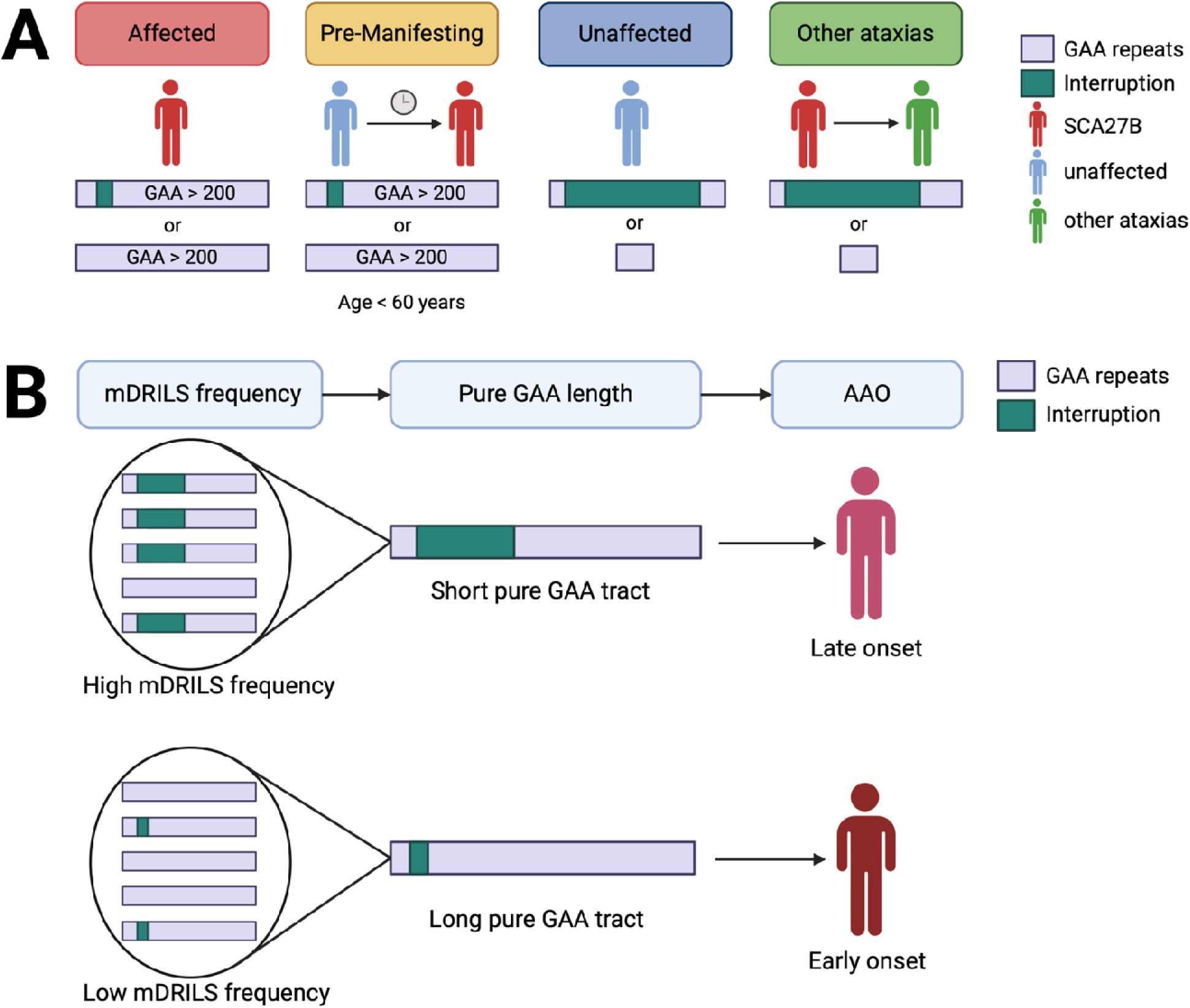
Schematic illustration of: (A) New categorization of individuals based on pure GAA length. Affected individuals with SCA27B, represented in red, have either >200 pure GAA repeats (i.e., without interruption) or >200 pure GAA repeats despite an interruption. Pre-manifesting carriers with an earlier age (<60 years of age) are highlighted in yellow. Unaffected individuals have long repeat interruptions and short pure GAA (<200 repeats) tracts. Lastly, misdiagnoses of SCA27B ataxia can occur in case of a long repeat interruption resulting in uninterrupted GAA repeats of < 200. **(B) Relationship between mDRILS frequency, pure GAA length, and age at onset.** mDRILS are modifiers of repeat stability and are more frequent in late-onset SCA27B compared to early-onset SCA27B. Legend: mDRILS = mosaic divergent repeat interruptions affection length and sequence, AAO = age at onset. Created in BioRender. Klein, C. (2025) https://BioRender.com/e06b901.

**Figure 4.**
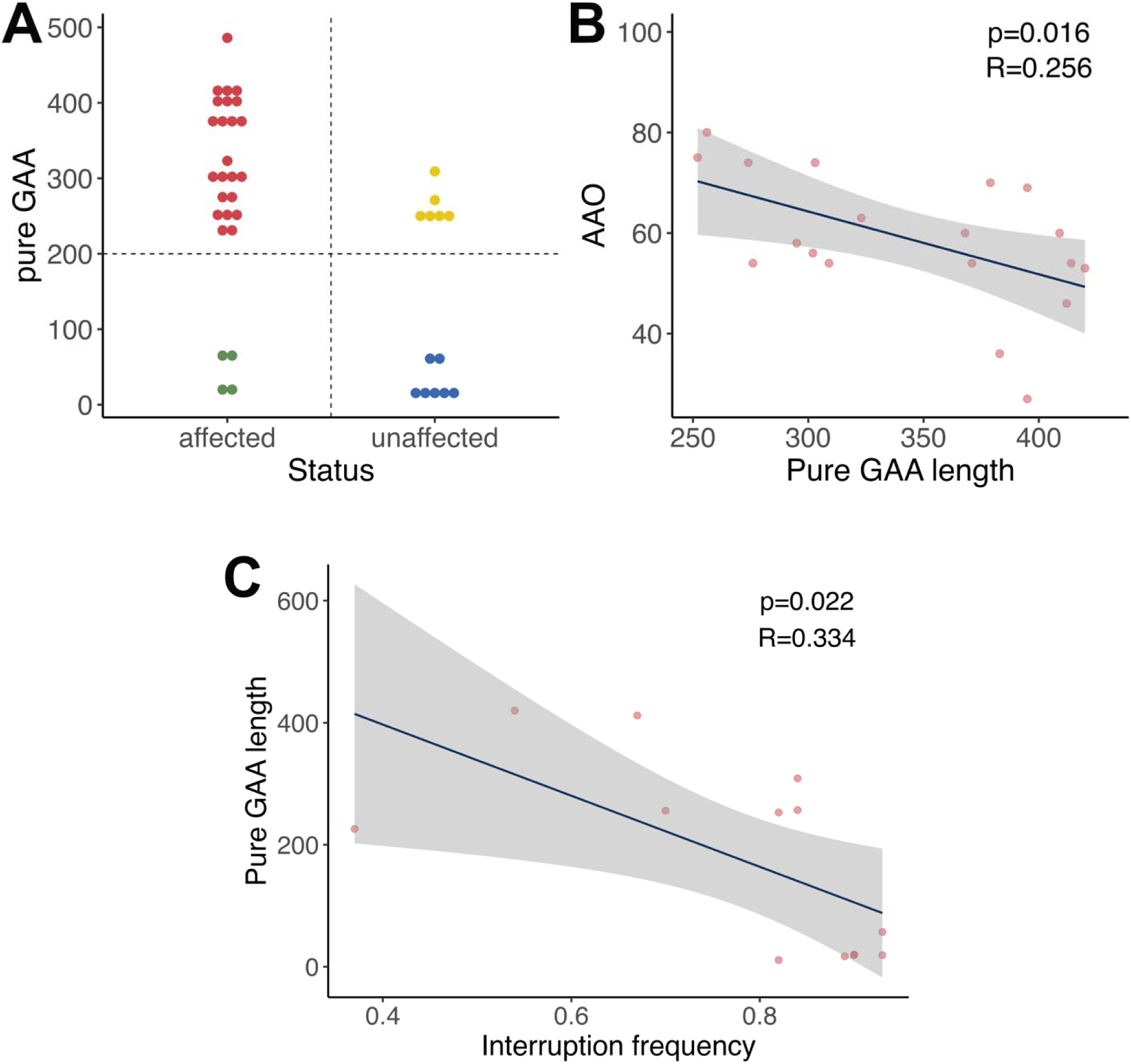
Analysis of the pure GAA length, determined by Nanopore sequencing. **(A)** Differences between affected and unaffected repeat expansion carriers in pure GAA length. Colour coding as described in Figure 3A. (i.e., red = affected, yellow = pre-manifesting, blue = unaffected, green = other ataxias) **(B)** Correlation between age at onset (AAO) and pure GAA length. **(C)** Correlation between interruption frequency and pure GAA length. A linear regression model was used for statistical analysis. The adjusted significance level is α= 0.013.

### Novel repeat interruptions were found through long-read sequencing

Seven different repeat interruptions were detected (Table 2). Among these, the most frequently observed interruption motif was (GAA*<u>GGA</u>*)_n_, characterized by a single adenosine to guanine conversion (A-to-*G*). This interruption motif was found in twelve individuals (six affected and six unaffected). The (GAA*<u>GGA</u>*)_n_ interruption motif was repeated two to 158 times and was located at the 5’end of the repeat tract, (GAA)_1-20_(GAA*<u>GGA</u>*)_2-158_ (GAA)_n_.

**Table 1:** Overview of the unaffected individuals with an expanded *FGF14* repeat expansion.

| ID | Status | Previously Published (PMID) | RN (ONT) | RN (LR-PCR) | RN (PacBio) | Interruption frequency (PacBio) | Pure GAA | Interruption in the long allele (ONT) | Methods confirming interruption |
| --- | --- | --- | --- | --- | --- | --- | --- | --- | --- |
| L-3013 | Unaffected | 36493768 | 316 | 326 | 318 | 0.90 | 20 | <u>GAAGGA</u> | PacBio, Sanger |
| L-3329 | Unaffected | 36493768 | 271 | 286 | 276 | - | 271 | - | Sanger |
| L-3344 | Unaffected | 36493768 | 319 | 326 | 318 | 0.90 | 19 | <u>GAAGGA</u> | PacBio, Sanger |
| L-3479 | Unaffected | 36493768 | 260 | 322 | 259 | 0.93 | 57 | <u>GAAGGA</u> | PacBio, Sanger |
| L-3501 | Unaffected | 36493768 | 319 | 326 | 318 | 0.93 | 19 | <u>GAAGGA</u> | PacBio, Sanger |
| L-3656 | Unaffected | 36493768 | 294 | 309 | - | - | 11 | <u>GAAGGA</u> | - |
| L-3657 | Unaffected | 36493768 | 309 | 326 | - | - | 309 | - | Sanger |
| L-8761 | Unaffected | 36493768 | 264 | 259 | 270 | 0.84 | 257 | <u>GAAAGAAGAAGGAA</u> | PacBio, Sanger |
| L-8782 | Unaffected | 36493768 | 256 | 256 | - | - | 256 | - | - |
| L-8934 | Unaffected | 36493768 | 315 | 289 | 314 | 0.89 | 17 | <u>GAAGGA</u> | PacBio, Sanger |
| L-10408 | Unaffected | 36493768 | 263 | 282 | 266 | 0.82 | 253 | <u>GAAAGAAGAAGGAAGAAGGAA</u> | PacBio, Sanger |
| L-10410 | Unaffected | 36493768 | 281 | 292 | - | - | 243 | <u>GAAAGAAGAAGGAAGAAGGAA</u> | Sanger |
| L-14996 | Unaffected | 37246629 | 309 | 309 | - | - | 66 | <u>GCA</u>GAAAGAAGAAGAA | - |

Legend: RN = repeat number, ONT = Oxford Nanopore Technology sequencing, LR-PCR = long-range PCR, PacBio = PacBio sequencing, Sanger = Sanger sequencing. Repeat motif variations are indicated by the underlined and italicized nucleotides.

**Table 2:**
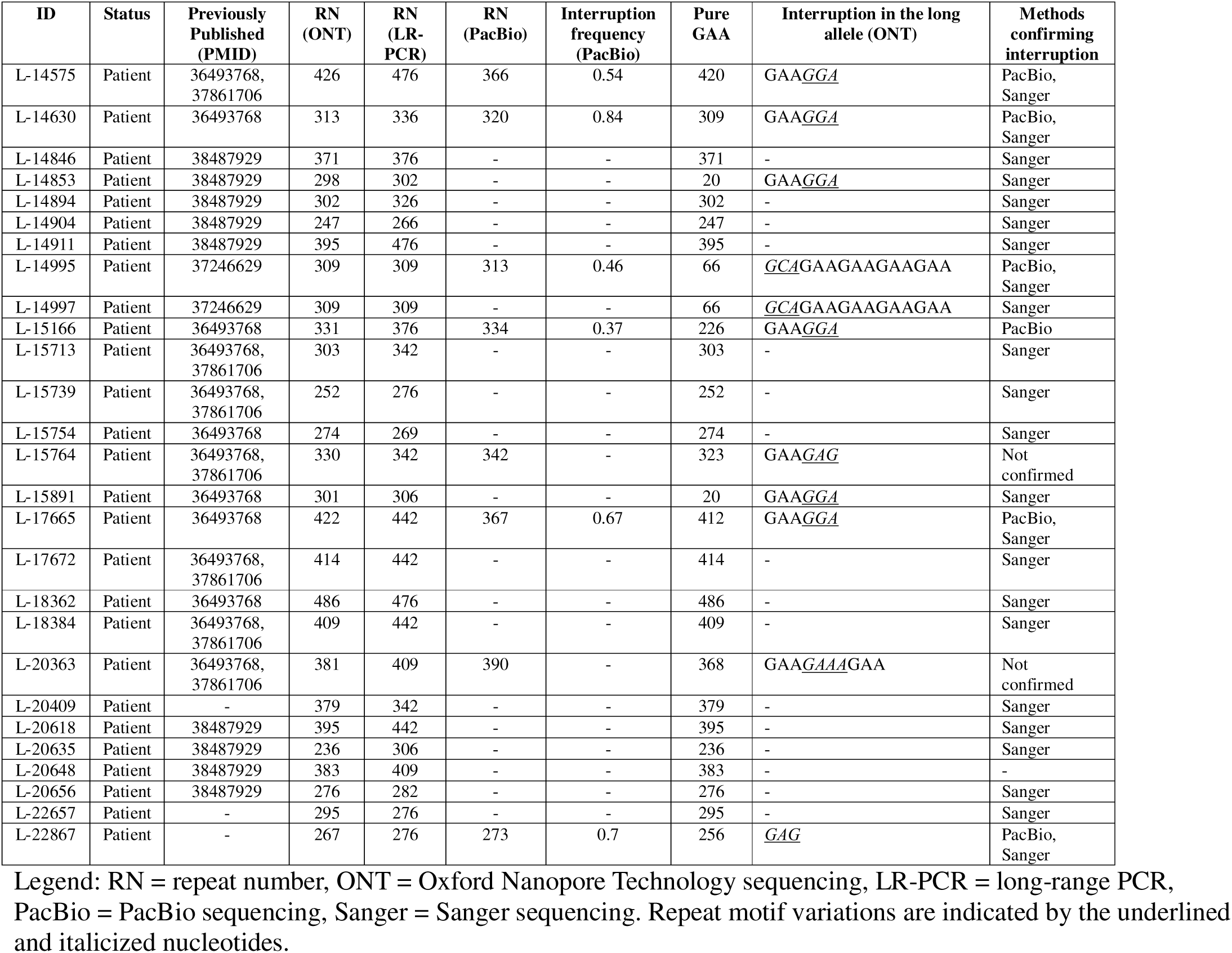
Overview of the patients with an expanded *FGF14* repeat expansion.

Another variant repeat motif was (GAA)_3_GAAA*<u>A</u>*GAAGAA*<u>G</u>*GAAGAA*<u>G</u>*GAA(GAA)_n_. This motif was identified in two unaffected individuals and included one deletion and two insertions. A similar motif, (GAA)_3_*G*AAA*<u>A</u>*GAAGAA*<u>G</u>*GAA(GAA)_n_, was identified in another unaffected. The shortest interruption motif observed was (GAA)_7_(*<u>GAG</u>*)_3_(GAA)_n_, detected in one patient.

Two motifs were exclusively detected by Nanopore sequencing. One motif was (GAA)_5_(GAAGAA*<u>A</u>*GAA)_3_(GAA)_n,_ which resulted from an insertion of an adenosine. The other motif was (GAA)_4_(GAA*<u>GAG</u>*)_2_(GAA)_n_. Both motifs were detected in patients.

The most complex motif was (GAA)_22_(*<u>GCA</u>*GAAGAAGAA)_3_(*<u>GCA</u>*GAA)(*<u>GCA</u>*GAAGAAGAAGAA)_24_(*<u>GCA</u>*GAA)_12_ (*<u>GCAGCA</u>*GAA)_3_(*<u>GCA</u>*GAA)_13_(*<u>GCAGCA</u>*GAA*<u>GCA</u>*GAA)_4_(*<u>GCA</u>*GAAGAAGAAGAA)_6_ (GAA)_22_*<u>GAG</u>*(GAA)_n_. This motif was observed in two affected members and one unaffected member of one family. However, the affected members of this family have a phenotype that differs from reported *FGF14* SCA27B patients.^4^

### Mosaic divergent repeat interruptions are associated with the pure GAA stability

Given the detection of mDRILS in XDP, we investigated the mosaicism of *FGF14* repeat interruptions in the long-read data. We calculated the mosaic frequency of the repeat motifs for each individual. The mosaicism obtained from Nanopore sequencing was similar to that from PacBio sequencing (*p*=0.017, *R*^2^=0.364) (Fig. 5A,B). We utilized PacBio sequencing data for mosaic frequency calculations due to its higher q-score. Notably, the repeat interruption mosaicism was negatively associated with pure GAA length (*p*=0.022, *R*^2^=0.334) (Fig. 4C). The higher the mosaic frequency, the shorter the pure GAA length.

**Figure 5.**
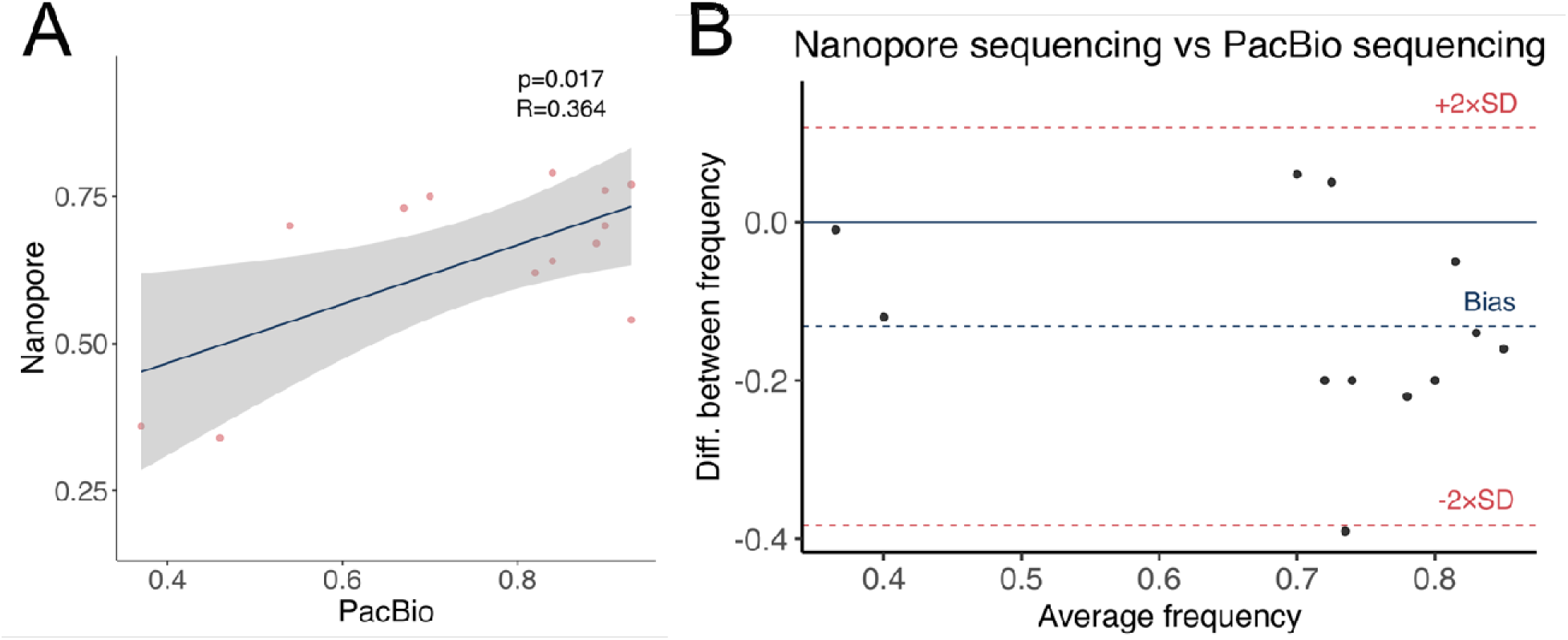
Comparison of the interruption frequency of all motifs between Nanopore and PacBio sequencing. **(A)** Correlation between Nanopore and PacBio sequencing. **(B)** Bland-Altman plot between Nanopore and PacBio sequencing. A linear regression model was used for statistical analysis.

To compare the interruption frequency between unaffected individuals and patients, we only included patients with the SCA27B phenotype; thus, a previously published Chilean family with a different phenotype was excluded.^20^ Unaffected individuals had higher interruption frequencies (0.90, IQR:0.89-0.90) compared to patients (0.67, IQR:0.54-0.70) (p=0.009, Z=-2.627) (Fig. 6A). The most common interruption motif (GAA*<u>GGA</u>*)_n_ had a mosaic frequency ranging from 0.37 to 0.93. Patients with (GAA*<u>GGA</u>*)_n_ exhibited a lower median frequency of 0.61 (IQR:0.50-0.71) compared to unaffected individuals, who had a median frequency of 0.90 (IQR:0.90-0.93) (*p*=0.014, *Z*=-2.470) (Fig. 6B). The mosaic frequencies of the GAAA*<u>A</u>*GAAGAA*<u>G</u>*GAAGAA*<u>G</u>*GAA motif and the GAAA*<u>A</u>*GAAGAA*<u>G</u>*GAA motif were similar, at 0.82 and 0.84, respectively. The shortest interruption motif, *<u>GAG</u>,* had a slightly lower frequency of 0.70 compared to the other motifs. In contrast to the other motifs, the most complex motif had the overall lowest frequency with 0.46.

**Figure 6.**
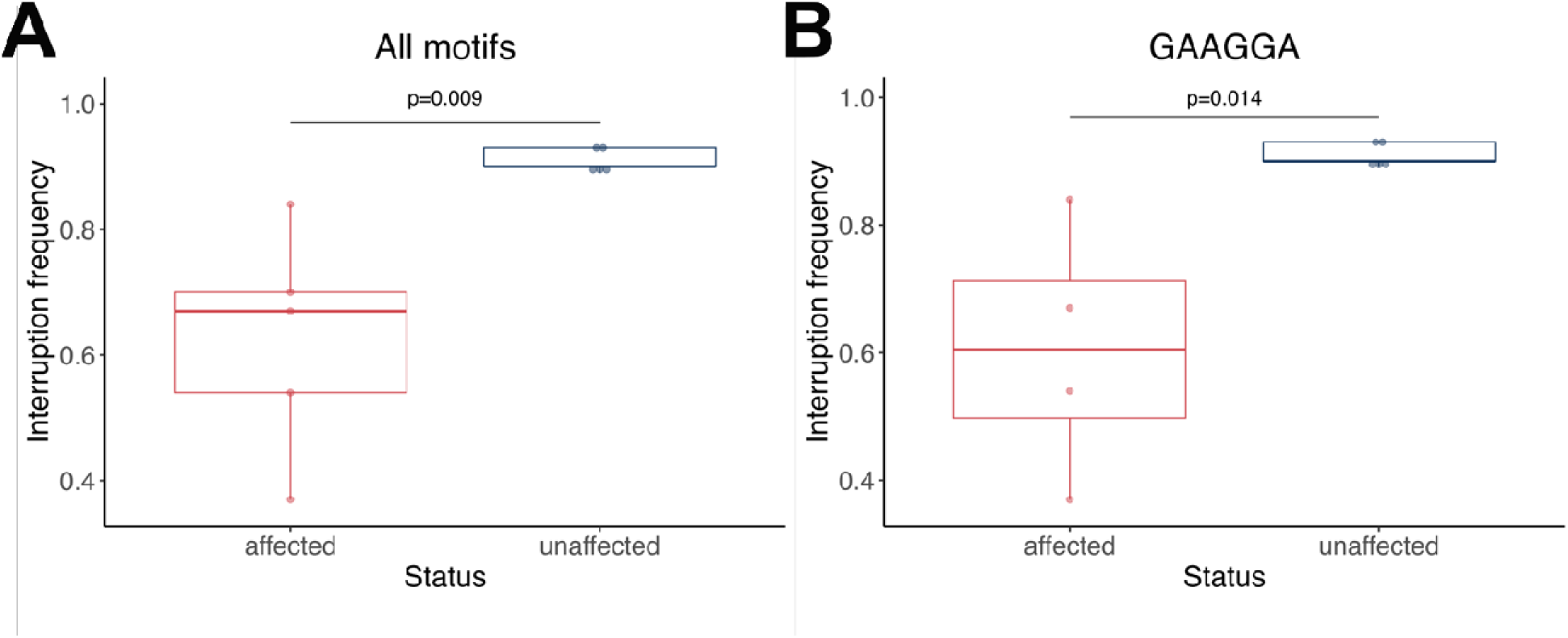
Analysis of the interruption frequency. Differences between affected and unaffected individuals in interruption frequency of all motifs **(A)** and the (GAA*<u>GGA</u>*)_n_ motif **(B)**. Mann-Whitney-U-tests were performed for statistical analysis. The adjusted significance level is α= 0.013.

Further investigation of repeat interruptions was performed to rule out DNA contamination from the non-expanded short allele. The median repeat number on the short allele was 18 (IQR:17-19). Repeat interruptions on the short allele were seen in three individuals (two patients, and one unaffected) (Supplementary Table 4). However, the interruption motifs in the short allele did not overlap with the expanded allele patterns.

While some interrupting sequences compromise the stability or toxicity of GAA repeats, others may stabilize the repeat or exert more severe damaging effects. Thus, we investigated the 5’-flanking the short and long allele in individuals with an interruption. All 20 individuals had a CTTTCTGT motif in the 5’-flanking region of the repeat motif on the long allele. For the short allele, three different motifs were identified. In ten of the 20 individuals with an interruption, the motif CTTTCTGTGTAGTCATAGTACCCC was detected. The motif CTTTCTGTG was identified in nine individuals. The third motif, CTTTCTGAG, was found in one individual. The 5’-flanking region of the short or long allele did not correlate with complex motifs nor interruption frequencies.

## Discussion

Following our discovery of mDRILS in X-linked dystonia-parkinsonism,^14^ we now observe that mDRILS can also act as modifiers of penetrance and age of onset (AAO) in a much more common condition, SCA27B. We consider this an important finding of translational relevance, as it suggests a novel, shared mechanism across different repeat expansion disorders, that has the potential to predict disease manifestation in individual repeat expansion carriers in a personalized fashion and represents a somatic phenomenon that may be amenable to environmental and lifestyle changes.

For the analysis of *FGF14* intronic repeat expansions, we propose a new concept to investigate and interpret their potential pathogenicity and challenge the previously held, more simplistic view of repeat interruptions, abolishing the pathogenic effect of the expanded repeat in general. Specifically, we propose four different scenarios, all related to the length of the pure GAA repeat tract: first, people with ataxia and an uninterrupted *FGF14* intronic repeat length of >200 repeats have SCA27B, despite the presence of an interrupted repeat, typically at the 5’ end of the expansion. All the patients in this group were examined by an experienced ataxia/movement disorder specialist and had a clinical syndrome consistent with the established SCA27B phenotype. Second, people with the same molecular constellation and age <60 years are considered to be in their premanifesting phase of SCA27B. Third, people with repeat interruptions resulting in a pure GAA repeat tract of <200 will not go on to develop SCA27B. Fourth, in patients previously presumed with SCA27B and carrying a large repeat interruption with <200 uninterrupted GAA repeats, the diagnosis needs to be revisited and likely revised to another type of ataxia (Fig. 2A). An example is the Chilean family with two members previously diagnosed with SCA27B but with a phenotype that differs from reported *FGF14* SCA27B patients,^4^ where we have identified a pure GAA tract with 65 repeat units. The threshold of 200 uninterrupted GAA repeats is a suggestion based on the data from the present study, albeit limited by our relatively low sample size. Therefore, a larger sample size is needed to validate and refine the real threshold of pure GAA repeats. Of further note, we here only included individuals with ≥250 repeats using long-range PCR, which was the starting point of our initial study based on the state of the literature at that point in time. However, our present data suggest that individuals carrying repeat expansions in the range of 200-250 repeats warrant further investigation, as this group may also harbor uninterrupted GAA repeat stretches of ∼200 repeats in length. This may further significantly broaden the group of *FGF14* GAA repeat expansion carriers who will likely manifest disease but are currently not considered to be at risk, according to present published recommendations.

Otherwise, our data implies that not all interruptions can be considered non-pathogenic, as, for example, five ataxia patients with interruptions still had a remaining pure GAA expansion >200. As many studies screened in unspecified ataxia cohorts for repeat expansions in *FGF14* to debunk the clinical phenotype of SCA27B without considering interruptions, our data demonstrates that 14% (4/28) of our ataxia patients with *FGF14* repeat expansions had rather non-pathogenic expansions if the presence of mDRILS is taken into account. Consideration of the pure GAA repeat tract and mDRILS is required, especially if the phenotype is not entirely consistent (e.g., early-onset disease, additional movement disorder features) with the typical clinical picture of SCA27B (i.e., pure late-onset cerebellar ataxia +/-episodic features). Moreover, the accurate detection of pure GAA repeats enabled by advanced sequencing methods such as Nanopore and PacBio sequencing might allow more robust genotype-phenotype relations, not only regarding the age at onset but also in the context of disease severity and progression. We suggest that it is feasible to utilize long-read sequencing in *FGF14* repeat expansion disorders in future clinical practice.

Notably, this situation in SCA27B extends beyond our previous findings in X-linked dystonia-parkinsonism, where the pathogenic insertion in the *TAF1* gene is the clear-cut cause in all affected individuals, and mDRILS in the hexanucleotide repeats within this insertion act as a modifier of AAO only. Intriguingly, in addition to the penetrance-determining length of the pure GAA tract in SCA27B, we can assume a similar AAO-modifying effect by mDRILS in *FGF14* as well, with a higher mDRILS frequency resulting in shorter uninterrupted GAA tracts, which, in turn, are associated with a later AAO (Fig. 3B). As a relationship between repeat length and age at onset is well-established in the field of repeat expansion disorders and, therefore, is expected to occur likewise in *FGF14*-related disease, our study shows the importance of taking into account mDRILS to better detect phenotypic correlations and impact on repeat stability.^15^ The mDRILS in both the XDP-relevant and *FGF14* repeats may affect repeat instability by modifying the propensity to form unusual mutagenic DNA structures, as observed with the interruptions of *FMR1* (FXS) and *ATXN1* (SCA1).^29^ As with other disease-associated repeat expansions, including the GAA expansions causing Friedreich’s ataxia,^10,30^ SCA27B-associated *FGF14* repeat expansions arise on specific haplotypes and can have interruptions in the repeats, suggesting the contribution of *cis*-elements both flanking and within the unstable repeat, respectively.^1,11,31,32^ There is a possibility that a similar mechanism and consequence of interrupting mDRILS play a role in Friedreich’s ataxia. However, long-read sequencing on repeat interruption motifs in Friedrich’s ataxia has not yet been performed. Evidence suggests an association of specific repeat interruptions with specific haplotypes (flanking *cis*-elements) that may predispose to parent-to-offspring (germline) repeat expansions.^1,11,31,32^ Possible mechanistic paths by which such flanking sequence changes may act, which may involve altered predisposition to variable chromatic impact and/or unusual nucleic acid structure formation.^11,31,32^

Importantly, the repeat sequence variations in *FGF14* we report herein arise somatically in post-natal tissues. We did not observe an association of the motif within the 5’-flanking region of the short allele and interruption motifs. Though our sample size is small and a larger number of individuals with interruptions are needed to thoroughly examine this. Experimental *in vivo* elucidation of the mechanism by which an intra-repeat or a flanking *cis*-element may act to modulate repeat instability, germline or somatic instability, can be quite complex. Animal studies of the SCA7 CAG tract,^33,34^ *in vitro* studies of the interrupted versus pure repeats of the SCA1/*ATXN1*, SCA10/*ATXN10*, and FXS/*FMR1* loci,^35–37^ and recent analysis of post-mortem tissues of the DM1/*DMPK*, *C9orf72,* and *FGF14* locus suggest chromatin compaction differences may mediate the function of *cis*-elements upon repeat instability.^11,38,39^

mDRILS-mediated alterations of the secondary and tertiary structures assumed by the repeat-containing mRNA may alter RNA-foci formation and can alter the proteins that may be bound and possibly sequestered from their normal functions. It is notable that the cerebellar ataxia, neuropathy, and vestibular areflexia syndrome (CANVAS)-associated (AAGGG)_n_•(CCCTT)_n_, but not the non-pathogenic expanded repeat motifs, including (AAAAG)_n_•(CTTTT)_n_, in *RFC1* can assume both triplex and quadruplex structures.^40–44^ The formation of these structures highlights the pathogenicity of the repeat sequence and can alter how the repeats are metabolized (replication/repair),^43^ which proteins can be bound and reveal novel manners by which they may be therapeutically targeted, for example, these pathogenic structures can be bound by the ligands TMPyP4 and BRACO-19.^40,43^ Somatically incurred mDRILS of the pathogenic *FGF14* repeat may also lead to pathogenic variations of spliceoforms, translation (exonization), repeat-associated non-AUG (RAN) translation, ribosomal frameshifting, ribosomal pausing, transcriptional slippage in the repeat, or repeat instability.^14,45–50^

The assessment of the *FGF14* mDRILS across time and generations would be beneficial to elucidate the potential impact of somatic changes in the mDRILS frequency on disease progression or severity. Longitudinal monitoring of patients with SCA27B and their mDRILS frequency was not performed in this study. However, our group previously investigated the mDRILS frequency in families affected by XDP, providing evidence that a higher frequency may stabilize the repeats across generations.^51^ Thus, future studies should aim for an analogous exploration of *FGF14* mDRILS across time within individual patients and across generations.

Overall, all methods (RP-PCR, Sanger sequencing, Nanopore sequencing, and PacBio sequencing) were concordant and detected the same interruptions. We confidently identified five out of seven different repeat interruption motifs within *FGF14.* Nanopore sequencing analysis performed thus far in *FGF14*-related disease could demonstrate that interruptions are present in both unaffected and ataxia patients but have not yet explored 1) the interruptions’ position and 2) their impact on repeat expansion pathogenicity at the individual level.^6^

Limitations of our study include the relatively small sample size compared to other studies. Moreover, we cannot independently support our estimate of pathogenicity based on advanced genetic methods, as we do not investigate *FGF14* expression, and further biomarkers robustly distinguishing SCA27B from other ataxias are not available to date. Thus, the development of such, preferably accessible biomarkers would be highly beneficial to the field. Future studies with larger sample size, longitudinal examinations, and more in-depth clinical data are needed to investigate not only the influence of pure GAA repeat number on age at onset but also a potential influence on disease severity and progression. Monitoring the mDRILS and their impact on disease progression and severity over a lifetime would be valuable. Finally, in the context of mosaicism, additional biomaterials such as fibroblasts, induced pluripotent stem cells, or post-mortem brain tissue should be investigated for mDRILS in future studies as well.

In conclusion, this study highlights the importance of an in-depth, multi-method assessment of repeat tract purity in repeat expansion disorders. The correlation of mosaic interruptions with the repeat stability and indirectly patient status suggests a protective effect of mDRILS. Long-read sequencing can uncover the length of the pure GAA motif along with repeat interruptions. However, it is warranted that the assessment of repeat interruptions, the pure GAA tract, and mDRILS be extended to other repeat expansion disorders and that their potential protective mechanism(s) be elucidated thoroughly. While our findings refine both the diagnosis of (*FGF14*-related) ataxia and the prognosis in non-manifesting or not-yet-manifesting repeat expansion carriers, it also highlights the necessity of including repeat interruption analysis not only in the research setting but also in diagnostic testing for SCA27B to avoid false-positive or false-negative testing results and interpretation thereof.

## Acknowledgment

We express our deepest gratitude to the families and patients who have participated in this study. We acknowledge Prof. Katja Lohmann for providing long-range and repeat-primed PCR analysis data of previously published and a small set of unpublished FGF14 expansion carriers and recognize her comments on the manuscript. We would also like to thank Frauke Hinrichs for her technical assistance.

## Supplementary material

Supplementary material is available online.

