## Supplementary for "*FGF14* repeat length and mosaic interruptions: modifiers of SCA27B?"

**Supplementary Table 1: Touchdown protocol for the long-range PCR**

| Temperature | Time | Cycles |
|---|---|---|
| 98°C | 1 min | |
| 98°C | 10 sec | 10x |
| 66°C | 10 sec | |
| 72°C | 2 min | |
| 98°C | 10 sec | 10x |
| 64°C | 10 sec | |
| 72°C | 2 min | |
| 98°C | 10 sec | 15x |
| 62°C | 10 sec | |
| 72°C | 2 min | |
| 72°C | 5 min | |

**Supplementary Table 2: Cycling conditions of the Repeat-primed PCR**

| Temperature | Time | Cycles |
|---|---|---|
| 98°C | 1 min | |
| 98°C | 10 sec | 10x |
| 64°C - 0.4/cycle | 10 sec | |
| 72°C | 2 min | |
| 98°C | 10 sec | 25x |
| 60°C | 10 sec | |
| 72°C | 2 min | |
| 72°C | 10 min | |

**Supplementary Table 3: Overview of the n=40 individuals with a repeat number > 250.**

| | Affected | Unaffected |
|---|---|---|
| AAO (Median (IQR)) | 57 years (48;68) | - |
| AAS (Median (IQR)) | 70 years (57;75) | 52 years (50;57) |
| RN (ONT) (Median (IQR)) | 313 (296;389) | 294 (264;315) |
| RN (PacBio) (Median (IQR)) | 320 (313;334) | 314 (268;318) |
| RN (LR-PCR) (Median (IQR)) | 342 (304;426) | 309 (286;326) |
| Frequency (Median (IQR)) | 0.61 (0.50;0.72) | 0.90 (0.89;0.92) |
| Interruption length (Median (IQR)) | 5 (3;44) | 124 (12;148) |

Legend: AAO = age at onset, AAS = age at sampling, RN = repeat number, ONT = Oxford Nanopore Technology sequencing, PacBio = PacBio sequencing, LR-PCR = long-range PCR, IQR = interquartile range.

**Supplementary Table 4: Overview of the repeat expansion on the short allele in the PacBio sequencing.**

| ID | RN | Interruption motif |
|---|---|---|
| L-3013 | 18 | - |
| L-3329 | 11 | - |
| L-3344 | 19 | - |
| L-3479 | 19 | - |
| L-3501 | 19 | - |
| L-3656 | 25 | *GAAAA* |
| L-8761 | 17 | - |
| L-8934 | 17 | - |
| L-10408 | 12 | - |
| L-14575 | 17 | - |
| L-14630 | 10 | - |
| L-14995 | 41 | - |
| L-15166 | 10 | - |
| L-15764 | 80 | *GAG*GAA*GAG* |
| L-17665 | 18 | - |
| L-20363 | 47 | GAA*GAAA*GAA |
| L-22867 | 17 | - |

Legend: RN = repeat number. Repeat motif variations are indicated by the underlined and italicized nucleotides.
